# COVID-19 case definitions, diagnostic testing criteria, and surveillance across the pandemic’s 25 highest burden countries

**DOI:** 10.1101/2021.05.11.21257047

**Authors:** Amitabh B. Suthar, Sara Schubert, Julie Garon, Alexia Couture, Amy M. Brown, Sana Charania

## Abstract

**Objective:** We compared suspect, probable, and confirmed case definitions, as well as diagnostic testing criteria, used in the COVID-19 pandemic’s 25 highest burden countries to aid interpretation of global and national surveillance data.

**Methods:** We identified the COVID-19 pandemic’s 25 countries with the highest disease burden based on the number of cumulative reported cases to the World Health Organization (WHO) as of 1 October 2020. We searched official websites of these countries for suspect, probable, and confirmed case definitions. Given that confirmation of COVID-19 usually requires diagnostic testing, we also searched for diagnostic testing eligibility criteria in these countries. Extracted case definitions and testing criteria were managed in a database and analyzed in Microsoft Excel.

**Findings:** We identified suspect, probable, and confirmed case definitions in 96%, 64%, and 100% of countries, respectively. Testing criteria were identified in 100% of countries. 56% of identified countries followed WHO recommendations for using a combination of clinical and epidemiological criteria as part of the suspect case definition. 75% of identified countries followed WHO recommendations on using clinical, epidemiological, and diagnostic criteria for probable cases. 72% of countries followed WHO recommendations on using PCR testing for confirming a case of COVID-19. Finally, 64% of countries used testing eligibility criteria at least as permissive as WHO.

**Conclusion:** There is marked heterogeneity in who is eligible for testing in countries and how countries define a case of COVID-19. This affects the ability to compare burden, transmission, and response impact estimates derived from case surveillance data across countries.

## Background

Novel infectious pathogens can pose major challenges to global health and security. Tracking the geography, demographics, and suspected mode of transmission of these pathogens using a standardized case definition remains the foundation for surveillance. Severe acute respiratory syndrome coronavirus 2 (SARS-CoV-2), the virus that causes coronavirus disease 2019 (COVID-19), was first characterized in December 2019.^1^ By January 2020, the first national case definition was developed^2^ and the World Health Organization (WHO) declared novel coronavirus disease 2019 a public health emergency of international concern.^3^ WHO’s interim guidance for global COVID-19 surveillance, released on 31 January 2020, included a hierarchy of confirmed, probable and suspect case definitions.^4^ This guidance encouraged the use of all available clinical, epidemiological and laboratory evidence for case classification purposes. In addition, it noted that countries may need to adapt the case definitions to their unique epidemiological situation. Recommendations for testing suspected cases and widespread testing were included based on transmission intensity, number of cases, and available resources.

Both WHO and national case definitions have evolved over time with growing knowledge about COVID-19 etiology and the myriad of ways the disease manifests after infection.^4–6^ Early on, surveillance emphasized a travel history to Wuhan, China, where the initial outbreak occurred, and a narrowly defined set of symptoms. However, the virus rapidly spread to other provinces in China and then internationally, and reports of patients who either experienced new symptoms or remained asymptomatic increased.^7^ Confirming a COVID-19 case relies on diagnostic testing and, therefore, testing capacity has played an important role in countries’ COVID-19 surveillance efforts. The types of tests available have expanded to include molecular and antigen tests to detect the presence of the SARS-CoV-2 virus and serology tests to detect antibodies produced from previous SARS-CoV-2 infection.^8,9^ However, the availability of these tests and the resources needed to collect, handle, and process clinical specimens have varied widely across nations.^10^ Shortages of test kits and reagents and lack of laboratory capacity have forced many locales to make tough decisions about policies for testing.^11^

Differences in case definitions pose a challenge not only to detect the true number of cases within countries, but also to understand the global burden of disease and adequately respond to pandemics. While global guidelines have been developed for testing eligibility criteria and case definitions, these are usually reviewed at a national level and are subject to adaptation based on laboratory and health system considerations. Earlier evaluations of global COVID-19 case definitions do not reflect the latest changes to national case definitions and test eligibility criteria and do not target the full range of countries with the highest burden of disease.^12–14^ In this study, we analyze (1) national COVID-19 case definitions from the 25 countries with the largest number of reported COVID-19 cases as of 1 October 2020 (collectively representing approximately 85% of the global cases at that time) and (2) specific criteria used to determine eligibility for SARS-CoV-2 diagnostic testing. The implications of inter-country differences on ongoing efforts to understand global disease burden and control the pandemic are also discussed.

## Methods

### Design

The 25 countries with the highest burden of COVID-19 disease were identified from WHO COVID-19 cumulative case counts as of 1 October 2020.^15^ The surveillance case definitions and official testing policies were extracted from official government websites for the respective countries. In instances where definitions were not available on official government websites, definitions were extracted from personal communication with the U.S. Centers for Disease Control and Prevention (CDC) field staff.

To find these data, we searched government websites using the following key words: case definition, suspect case, confirmed case, COVID-19, case criteria, surveillance, testing criteria, guidelines, laboratory, reverse transcriptase-polymerase chain reaction (RT-PCR), and asymptomatic. All surveillance definitions and testing criteria were verified as current as of 1 January 2021. Several of the official policies were not available in English. For these documents, we used Google Translate to identify the definitions and testing policies.

### Data management and analysis

In order to compare case definitions across countries, we classified the components of each definition into three parts: (1) diagnostic components including a laboratory test or radiographic imagery; (2) clinical symptoms (such as cough, fever, and Severe Acute Respiratory Infection (SARI)); and (3) epidemiological criteria, including travel to a high-burden region or contact with a confirmed or suspect case. For each country’s testing policy, we considered individuals eligible for diagnostic testing. Countries were classified as testing asymptomatic individuals without any additional criteria, testing asymptomatic individuals with some epidemiological criteria, such as contact with a confirmed case, or recommending testing exclusively for symptomatic individuals. These analyses were based solely on diagnostic testing eligibility criteria and did not consider exceptions, such as testing asymptomatic individuals prior to travel, asymptomatic testing through the private sector, or local-level mass-testing. Elements of national case definitions and testing criteria were compared against global norms from WHO.

### Source assessment

To assess sources, we extracted information on their origin and timeliness. The origin was categorized as a government source or personal communication while timeliness was based on date of publication.

## Results

### Suspect case definitions

We identified suspect case definitions in 24 of 25 countries (96%) (Table 1 and Appendix 1). Israel relies on surveillance via cell phone data; we used these epidemiological criteria from Israel to create a suspect case definition. The three most common criteria included in suspect case definitions were fever, cough, and labored breathing (reported in 92%, 84%, and 84% of 25 countries). Seven countries (28%) used “other” criteria in addition to the common criteria listed in Table 1. The WHO suspected case definition includes clinical symptoms, including the three most common stated above, and epidemiological criteria. Fourteen (56%) countries followed this guidance broadly by including clinical and epidemiological criteria, ten (40%) countries required clinical symptoms alone for the suspect case definition, while two countries (8%) also incorporated diagnostic testing. The United States relies on laboratory evidence, including antibody or antigen positivity, without any clinical symptoms or epidemiological criteria while Colombia primarily relies on epidemiological criteria and clinical symptoms, but includes laboratory and radiological tests as part of their definition to assist with diagnoses.^16,17^

**Table 1.**
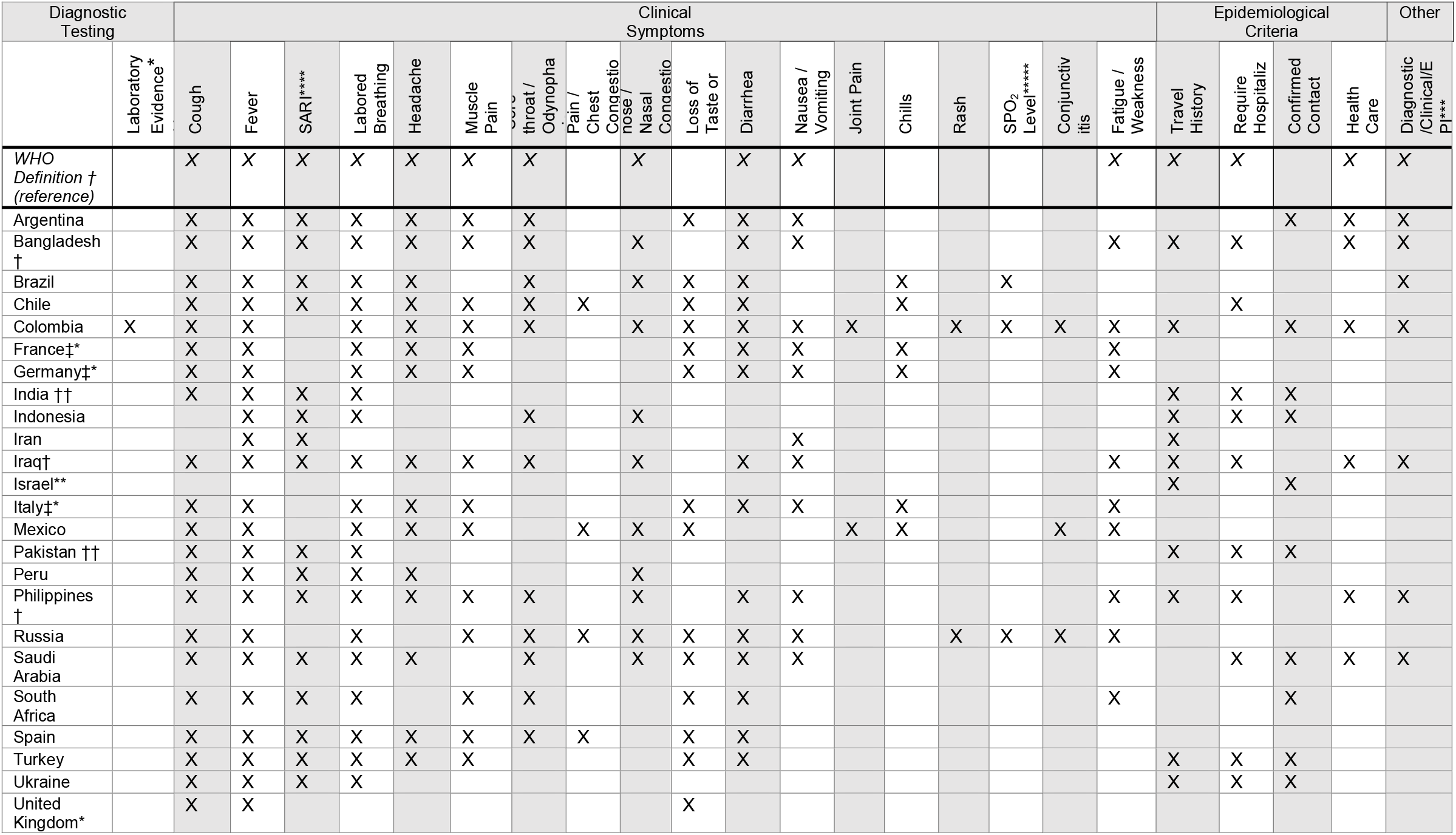
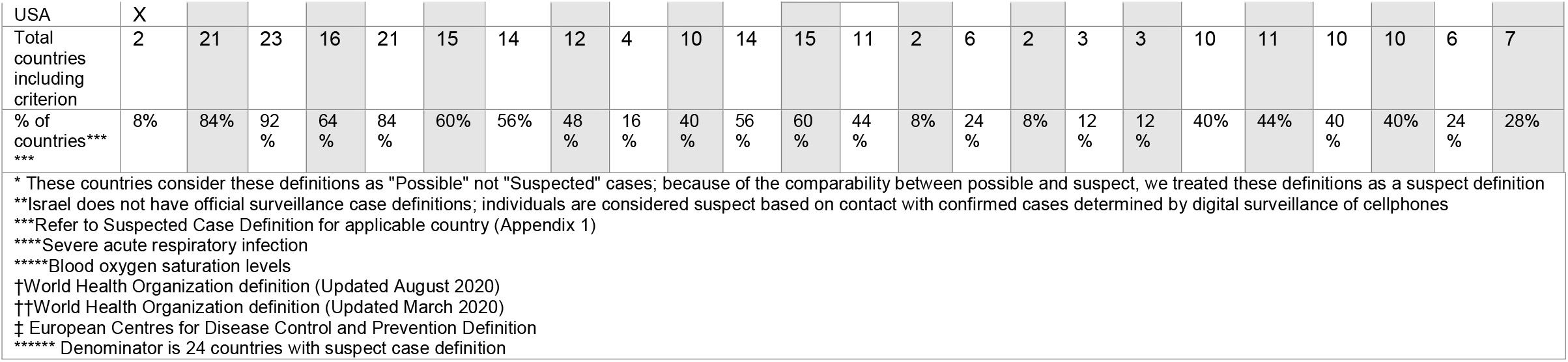
Suspect case definition criteria across the COVID-19 pandemic’s 25 highest burden countries, current as of 1 January 2021. ‘X’ indicates the criterion was sufficient for, or a potential component of, the suspect case definition requirement(s). Full suspect case definitions can be found in Appendix 1.

### Probable case definitions

We identified probable case definitions in 16 of 25 countries (64%) (Table 2 and Appendix 2). The WHO probable case definition includes criteria from all three categories, diagnostic testing (chest imagery), clinical symptoms, and epidemiological criteria. 12 out of 16 countries (75%) were consistent with WHO and included criteria from all three categories. There was heterogeneity in the number of required criteria across countries. The three most common criteria included in probable case definitions were fever, labored breathing, and confirmed contact with a probable or confirmed case (reported in 94%, 88%, and 81% of the 16 identified countries, respectively). Fourteen (88%) countries included some type of diagnostic testing for the probable case definition, fifteen (94%) included clinical symptoms in their definitions, and fourteen (88%) included epidemiological criteria.

**Table 2.**
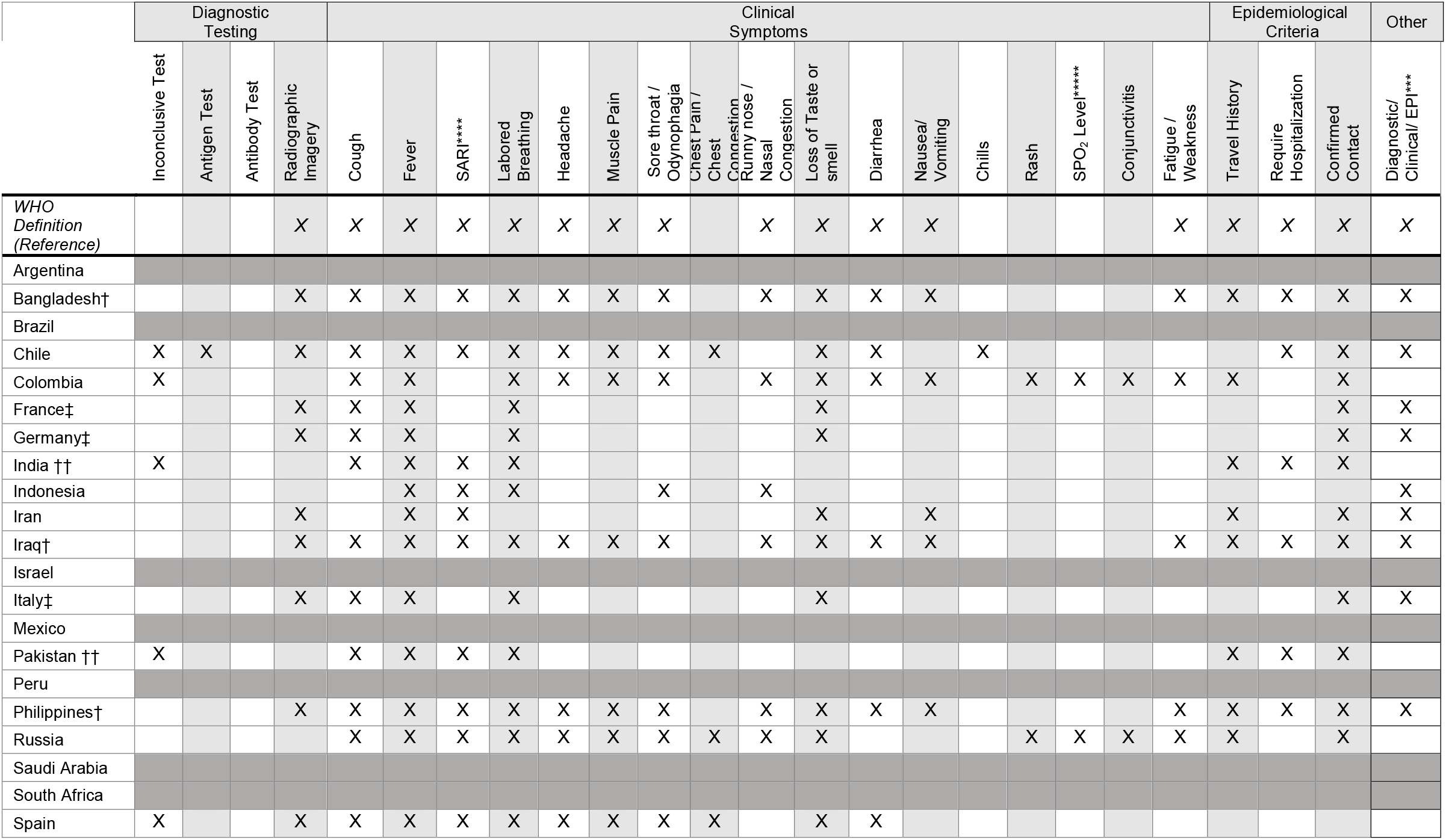
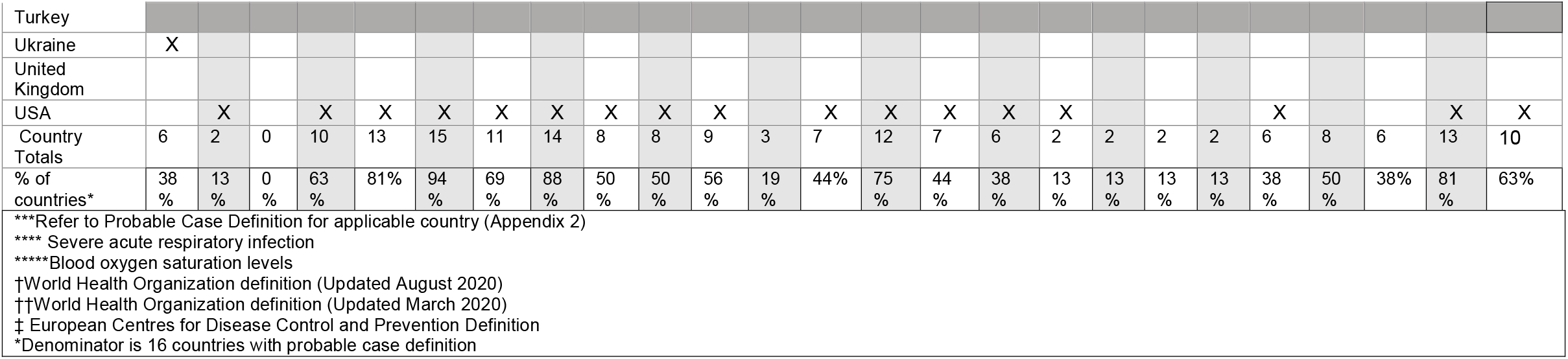
Probable case definition criteria across the COVID-19 pandemic’s 25 highest burden countries, current as of 1 January 2021. ‘X’ indicates the criterion was sufficient for, or a potential component of, the probable case definition requirement(s). Full probable case definitions can be found in Appendix 2.

### Confirmed case definitions

We identified confirmed case definitions in all 25 countries (100%) (Table 3 and Appendix 3). All confirmed case definitions required some type of diagnostic testing. Eighteen (72%) countries were consistent with WHO’s recommendations and specified RT-PCR tests in their case definition. Of these countries, ten (40%) also included antigen or antibody tests in their definition. Seven countries (28%) did not specify a type of diagnostic test. Seven countries (28%) include reference to their suspect case definition within their confirmed case definition. Of these, three countries (Mexico, Saudi Arabia, and Turkey) require that an individual meet the suspect case definition in addition to diagnostic testing criteria. In addition to confirming cases based on diagnostic testing, six (24%) countries confirmed cases exclusively on loss of taste or smell (anosmia or ageusia). Overall, eight countries (32%) included clinical symptoms as part of their confirmed case definition.

**Table 3.**
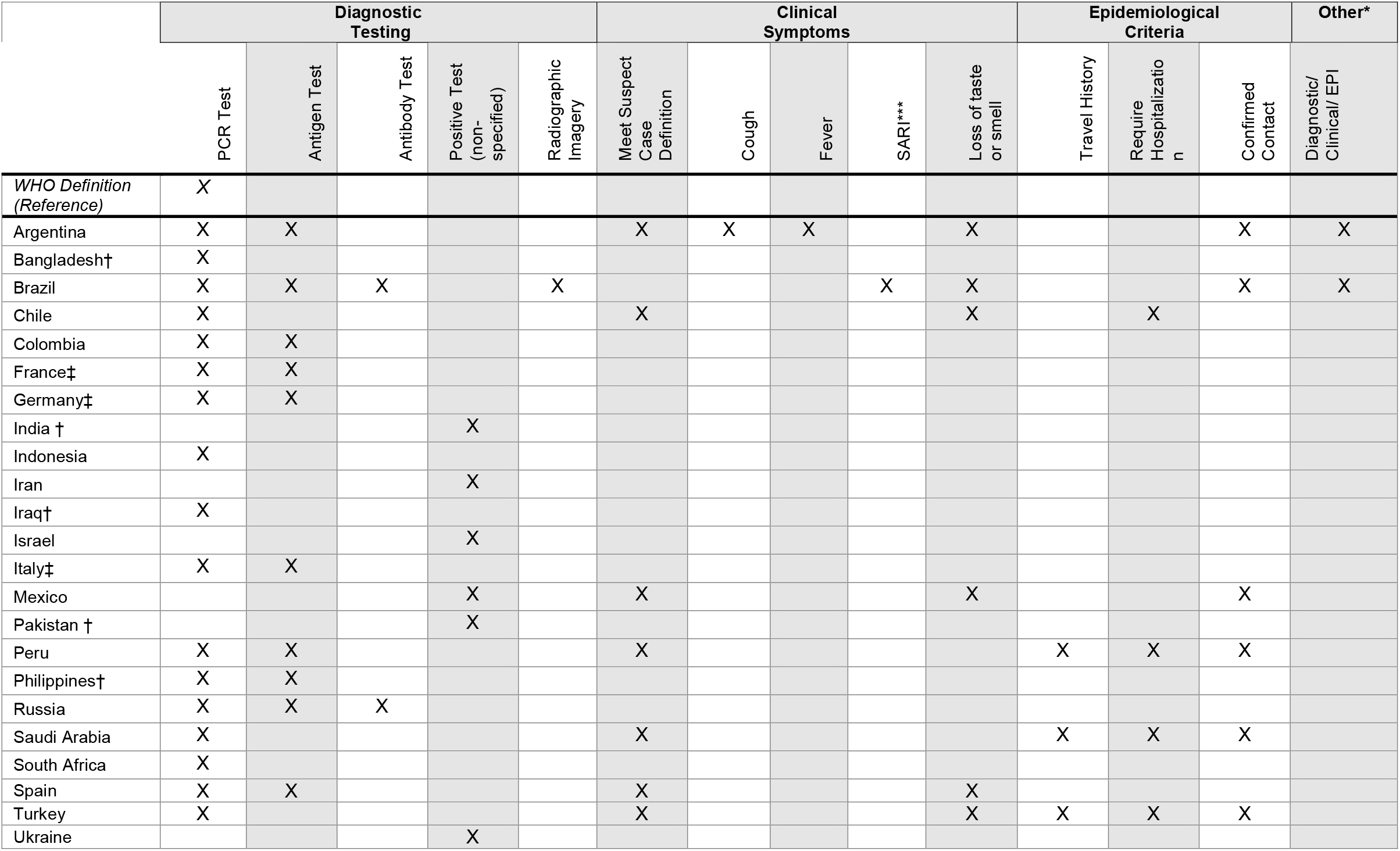
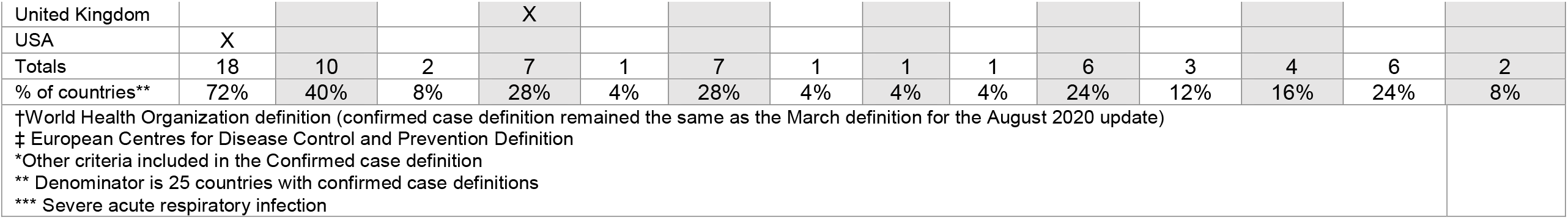
Confirmed case definition criteria across the COVID-19 pandemic’s 25 highest burden countries, current as of 1 January 2021. ‘X’ indicates the criterion was sufficient for, or a potential component of, the confirmed case definition requirement(s). Full confirmed case definitions can be found in Appendix 3.

### Testing eligibility criteria

We identified testing criteria in all 25 countries (100%) (Appendix 4). Eight (32%) of countries had no symptom requirements for testing, eight (32%) had no symptom requirements for testing but required epidemiological criteria, i.e., exposure to a confirmed or probable case, and nine (36%) countries required symptoms. Of the eight countries requiring epidemiological criteria, five (63%) also allowed asymptomatic testing for a healthcare worker (Appendix 4). Policies from Saudi Arabia and the United Kingdom specified not to test asymptomatic individuals but included an exception for healthcare employees (Appendix 4). WHO recommends testing asymptomatic individuals who have had contact with a confirmed case; 64% of countries used eligibility criteria at least as permissive as WHO.

### Source assessment

92% of case definitions were found on official government websites and 72% were published or included in documents published after the most recent WHO definition was published (7 August 2020) (Table 4). Two countries (India and Pakistan) utilized the previous WHO definition (dated March 2020). We were unable to confirm that these countries updated their definition based on the newest WHO definitions. We were unable to locate definitions on Israel, Iraq, and Iran’s government websites. In these countries, we obtained definitions from personnel involved in the respective country’s COVID-19 response. 23 of 25 countries (92%) had an official government source for diagnostic testing criteria. Iraq and Iran testing criteria were obtained via personal communication. 88% of testing criteria were published after 1 September 2020. The policies for Philippines, Brazil, and Pakistan were updated in July and August 2020.

**Table 4.** Assessment of sources for case definition and testing criteria across the COVID-19 pandemic’s 25 highest burden countries, current as of 1 January 2021.

| Countries | Case Definition Source (Date)* | Testing Criteria Source (Date)* |
| --- | --- | --- |
| WHO | WHO<sup>6</sup><br>(8/7/2020) | N/A |
| Argentina | Government<sup>34</sup><br>(9/11/2020) | Government<sup>35</sup><br>(9/23/2020) |
| Bangladesh | Government<sup>36</sup><br>(11/5/2020) | Government<sup>36</sup><br>(11/5/2020) |
| Brazil | Government<sup>37</sup><br>(8/5/2020) | Government<sup>37</sup><br>(8/5/2020) |
| Chile | Government<sup>38</sup><br>(10/1/2020) | Government<sup>39</sup><br>(11/18/2020)*** |
| Colombia | Government<sup>17</sup><br>(10/19/2020) | Government<sup>40</sup><br>(10/2020) |
| France | ECDC<sup>41</sup><br>(12/3/2020) | Government<sup>42</sup><br>(10/19/2020)*** |
| Germany | ECDC<sup>41</sup><br>(12/3/2020) | Government<sup>43</sup><br>(12/16/2020) |
| India | Government<sup>44</sup><br>(7/3/2020) | Government<sup>45</sup><br>(9/4/2020) |
| Indonesia | Government<sup>46</sup><br>(7/13/2020) | Government<sup>47</sup><br>(1/1/2021)** |
| Iran | CDC contact<sup>48</sup><br>(9/29/2020)* | CDC contact<sup>48</sup><br>(9/29/2020)* |
| Iraq | CDC contact<sup>49</sup><br>(10/2/2020)* | CDC contact<sup>49</sup><br>(10/2/2020)* |
| Israel | CDC contact<sup>50</sup><br>(10/7/2020)* | Government<sup>51</sup><br>(12/17/2020) |
| Italy | ECDC<sup>41</sup><br>(12/3/2020) | Government<sup>52</sup><br>10/23/2020) |
| Mexico | Government<sup>53,54</sup><br>(8/25/2020) | Government<sup>55</sup><br>(11/11/2020) |
| Pakistan | Government<sup>56</sup><br>(3/27/2020) | Government<sup>57,58</sup><br>(12/2/2020) |
| Peru | Government<sup>59</sup><br>(7/10/2020) | Government<sup>60</sup><br>(9/30/2020) |
| Philippines | Government<sup>61</sup><br>(11/25/2020) | Government<sup>62</sup><br>(7/6/2020) |
| Russia | Government<sup>63</sup><br>(10/26/2020) | Government<sup>64</sup><br>(1/1/2021)** |
| Saudi Arabia | Government<sup>65</sup><br>(10/2020) | Government<sup>65</sup><br>(10/2020) |
| South Africa | Government<sup>66</sup><br>(8/18/2020) | Government<sup>67</sup><br>(9/16/2020) |
| Spain | Government<sup>68</sup><br>(12/18/2020) | Government<sup>68</sup><br>(12/18/2020) |
| Turkey | Government<sup>69</sup><br>(12/7/2020) | Government<sup>69</sup><br>(12/7/2020) |
| Ukraine | Government<sup>70</sup><br>(3/28/2020) | Government<sup>71</sup><br>(1/1/2021)** |
| UK | Government<sup>72</sup><br>(9/28/2020) | Government<sup>73</sup><br>(1/1/2021)** |
| USA | Government<sup>16</sup><br>(8/5/2020) | Government<sup>8</sup><br>(10/21/2020) |

\* Date of U.S. Centers for Disease Control and Prevention (CDC) contact communication
\*\*Date the source website was last verified in absence of website update date
\*\*\* Date the website was last updated

## Discussion

A consistent statement across all iterations of WHO’s global COVID-19 surveillance guidance is that countries may need to adapt the case definitions to their specific circumstances.^4–6,18^ Beginning with the 20 March 2020 version, WHO also encouraged countries to publish their adapted versions online and in periodic situation reports.^5,6^ Nearly all countries (92%) in this analysis indeed chose to deviate from WHO’s case definitions in some manner and 92% of countries posted their case definition on an official government website. Suspect and confirmed case classifications were found for nearly all countries, but over a third (36%) excluded the probable case classification. In addition, substantial variation was observed among testing criteria used across the national case definitions. While WHO reserved the use of laboratory testing for confirmed cases only, two countries (8%) included laboratory evidence for suspect cases, 14 countries (88%) for probable cases, and nearly a third (32%) included non-laboratory criteria for confirmed cases. Laboratory evidence in some countries was not restricted to RT-PCR, but rather included increasingly available antigen and antibody tests. Lastly, testing eligibility criteria also differed widely with many countries either excluding asymptomatic individuals from routine testing (36%) or only including them under certain conditions (32%).

Differences in case definitions and testing eligibility can have important implications on efforts to monitor disease trends and understand the impact of vaccine scale-up efforts across countries and over different time periods. As knowledge of a novel disease increases, the sensitivity and specificity of the case definition changes over time, ultimately impacting the number of cases identified.^19^ For example, in 2003 during the earlier SARS epidemic, the Netherlands had several iterations of case definitions that diverged from the prevailing WHO case definition, which was more sensitive and less specific in comparison. When all cases were reevaluated, 21 cases were classified as suspect and two as probable using the latest WHO case definition as opposed to just nine suspect and zero probable cases using the Dutch case definitions.^20^ As the COVID-19 pandemic emerged in China, a 12 February 2020 case definition change to include clinically diagnosed mild cases resulted in identification of more than 15,000 cases in a single day.^21^ A study of China’s successive case definitions, each with gradually increasing sensitivity, also yielded higher detection of cases.^14^ From 15 January to 3 March 2020, China’s National Health Commission used seven versions of the case definition for COVID-19, estimated to increase the proportion of cases being detected by seven times after the first change, three times from change two to four, and four times after the fifth change. The authors estimated that if the fifth version of the case definition had been applied throughout the outbreak, there would have been 4.2 times more confirmed cases identified in China by 20 February 2020 (232,000 compared to 55,508). These data suggest that use of less sensitive case definitions can underestimate the burden of COVID-19. Changes in case definitions may need to be considered while analyzing an epidemic curve for COVID-19 or other novel diseases.

The wide variation we found in suspect and probable case criteria – and the complete omission of the probable case classification in some nations – is of particular interest. When test results are still pending or tests are unavailable, inclusion of suspected and probable cases allows for early isolation and treatment of these cases.^7^ In addition, in its 7 August 2020 guidance, WHO requested that countries include counts of probable cases along with confirmed cases in weekly aggregate reports.^6^ WHO indicated that suspected and confirmed case definitions were revised to reflect increased knowledge of the clinical spectrum of COVID-19 signs and symptoms, especially the most common and predictive. These updates were important to informing global and national surveillance because some symptoms have been found to have limited predictive value for surveillance purposes despite their frequent inclusion in case identification procedures.^22–25^

WHO case definition guidance does not explicitly state the type of test for diagnostic confirmatory testing but references the laboratory guidance that recommends nucleic acid amplification tests (NAAT), such RT-PCR assays.^26^ Many counties may not have considered the laboratory guidance and used the WHO confirmed case definition verbatim. Indeed, we found that seven of the high burden countries did not specify the type of test to be used in confirmatory testing. Results indicating some countries’ use of alternatives to NAAT as laboratory evidence is another important finding. Antigen tests, particularly point of care tests, have been promoted as an important tool for early detection and to prevent asymptomatic spread.^9^ However, their sensitivity is generally lower than NAAT leading to false negatives.^27^ Antibody tests have typically been recommended as a surveillance assay rather than a standalone diagnostic tool.^11,28^ Despite the limitations of NAAT alternatives, they are increasingly available in many areas and have important benefits, such as lower overall cost, simplified logistics and supply chain management, and faster turnaround of results for rapid versions that may explain their integration in some national confirmed case definitions.

WHO’s public health surveillance guidance also includes recommendations for laboratory testing of all suspect and probable cases but acknowledges that testing priorities would be dependent on intensity of transmission, the number of cases, and laboratory capacity. Therefore, guidance was also developed on testing asymptomatic and mildly symptomatic individuals.^6,26^ National differences in testing eligibility criteria may also reflect resource limitations and differences in national health insurance coverage of tests.^12^ A number of testing strategies proposed to target segments of the population believed to be at greatest risk of exposure to SARS-CoV-2. For example, some have proposed testing all symptomatic individuals and asymptomatic individuals with known or suspected contact with COVID-19 case for optimal NAAT testing.^9,26^ Testing asymptomatic individuals without an exposure was suggested if results would impact isolation, quarantine, personal protective equipment usage decisions, surgery eligibility, or inform administration of immunosuppressive therapy.^29^ In Australia, the national testing policy emphasized defining and targeting high-risk settings, such as residential care facilities or correctional facilities for testing.^30^ In May 2020, the European Centre for Disease Prevention and Control expanded the pool of individuals eligible for laboratory testing, resources permitting, to include asymptomatic individuals in healthcare settings and long-term care facilities, to identify potential sources of infection and protect vulnerable individuals.^13^

In order to get accurate case counts, detecting both symptomatic and asymptomatic cases is necessary due to the large proportion of COVID-19 cases presenting with no or mild symptoms.^29^ Inclusion of asymptomatic cases also impacts key epidemiological metrics, such as incidence and the case fatality ratio. While expansive testing criteria will increase the likelihood of capturing asymptomatic infections, they should be balanced against the burden for the public health system in tracing and testing these eligible individuals.^30^ For example, broadening testing eligibility criteria may overload the healthcare system with individuals with a low probability of infection and/or disease progression. Furthermore, many settings may not have enough resources to test all eligible individuals.^29^ Although WHO provides harmonious global testing criteria and case definitions, our findings suggest heterogeneity in how these were adapted and that it might be necessary to account for these deviations when comparing COVID-19 case trends across countries.

There are several methodological limitations to our analysis. First, although the included studies represented approximately 85% of the reported cases globally, there may be differences in case definitions among the countries comprising the remaining 15%. Second, although we identified suspect case definitions, confirmed case definitions, and testing criteria for most countries, we only identified probable case definitions in 16. This may be due to the lack of a probable case definition or the lack of making it available in the public domain; regardless, the results of the probable case definition analyses may be less generalizable than the others. Third, since Google Translate was used to translate definitions not in English, there may have been discrepancies between the original policy intent and what we were able to interpret from the translation. Fourth, our scope was limited to confirmed, probable and suspect case definitions; other classifications, such as persons under investigation, may merit further research. Fifth, after extraction and analyses were completed, additional issues relating to case surveillance have emerged. These include criteria for distinguishing a new case from an existing case, i.e., reinfection cases, as well as the B.1.1.7, 1.351, and P.1 variants of COVID-19.^16,31–33^ Although neither reinfection nor genomic sequencing were part of national case surveillance definitions, reinfection surveillance may provide further information on naturally and vaccine-acquired immunity whilst genomic surveillance may provide further insights on circulating strains and are vital elements of comprehensive national surveillance of COVID-19. Sixth, given the large number of possibilities, we chose not to list every permutation of laboratory, clinical, and epidemiological criteria for the WHO and national suspect, probable, and confirmed case definitions. Finally, despite our analysis of each government’s policies, these policies may not be implemented equally in various settings and may change over time.

Case surveillance remains the foundation for national COVID-19 surveillance and will play a vital role in understanding the impact of national scale-up of vaccines and continued mitigation efforts. There is marked heterogeneity in who is eligible for testing among countries and how countries define a case of COVID-19. Specifically, we observed heterogeneity in elements used across the suspect case definitions (heterogeneity in eligible clinical symptoms), probable case definitions (heterogeneity in laboratory and diagnostic requirements), and confirmed case definitions (with some countries using antigen or PCR and others only used PCR testing). Importantly, testing eligibility criteria varied across countries from being restricted to populations with exposure and symptoms to not having any requirements. Collectively, these issues suggest that efforts to compare trends of COVID-19 across countries require careful interpretation.

## Data Availability

All relevant data are available in the manuscript body and appendices.

## Appendix 1.

Suspect Definitions

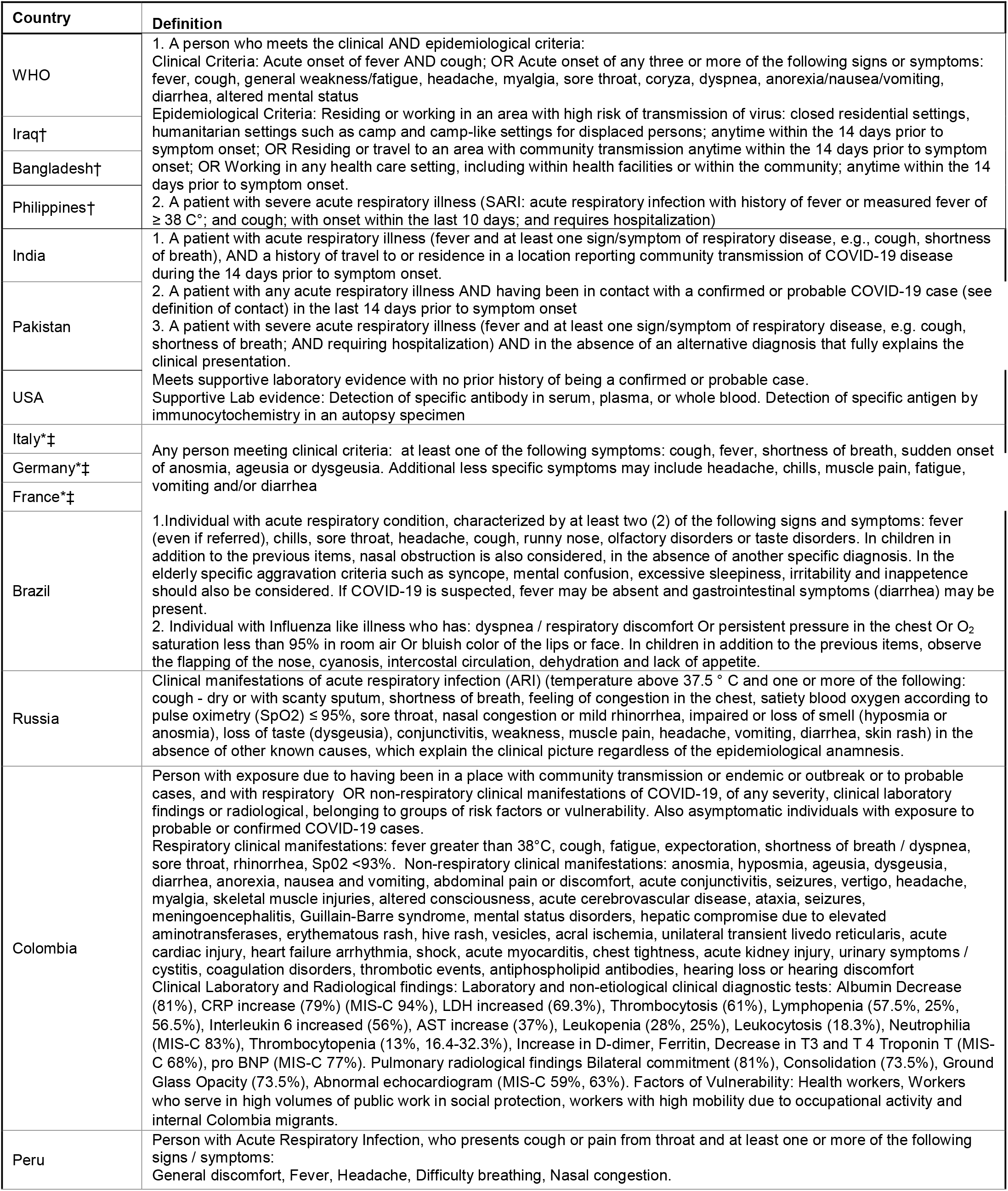
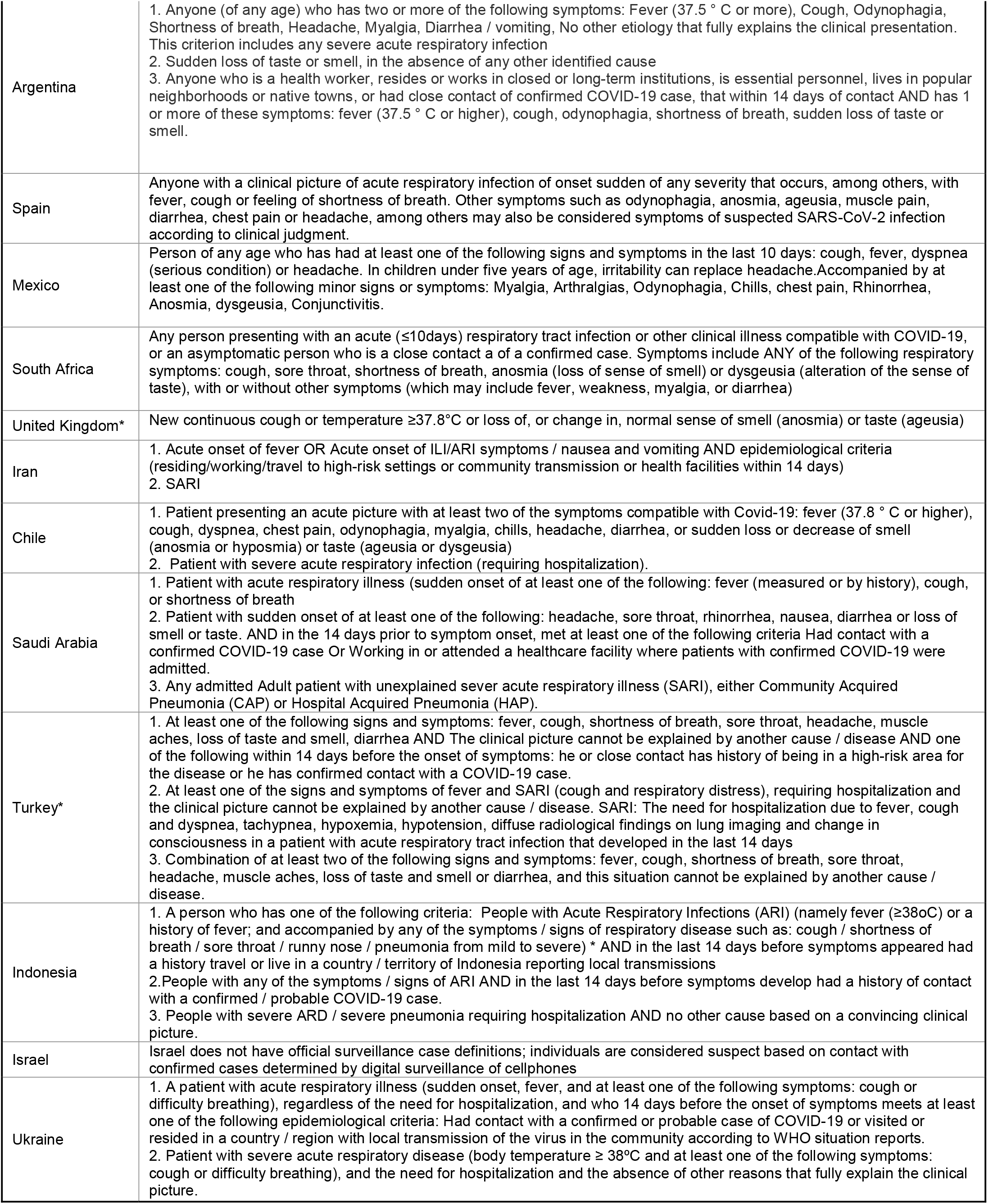
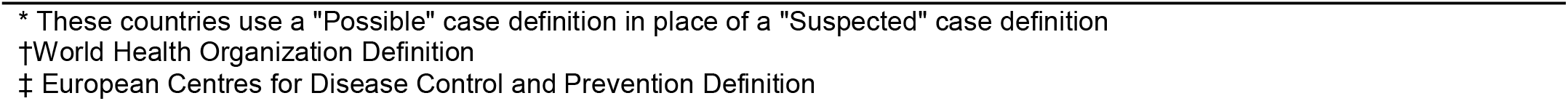

## Appendix 2.

Probable Definitions

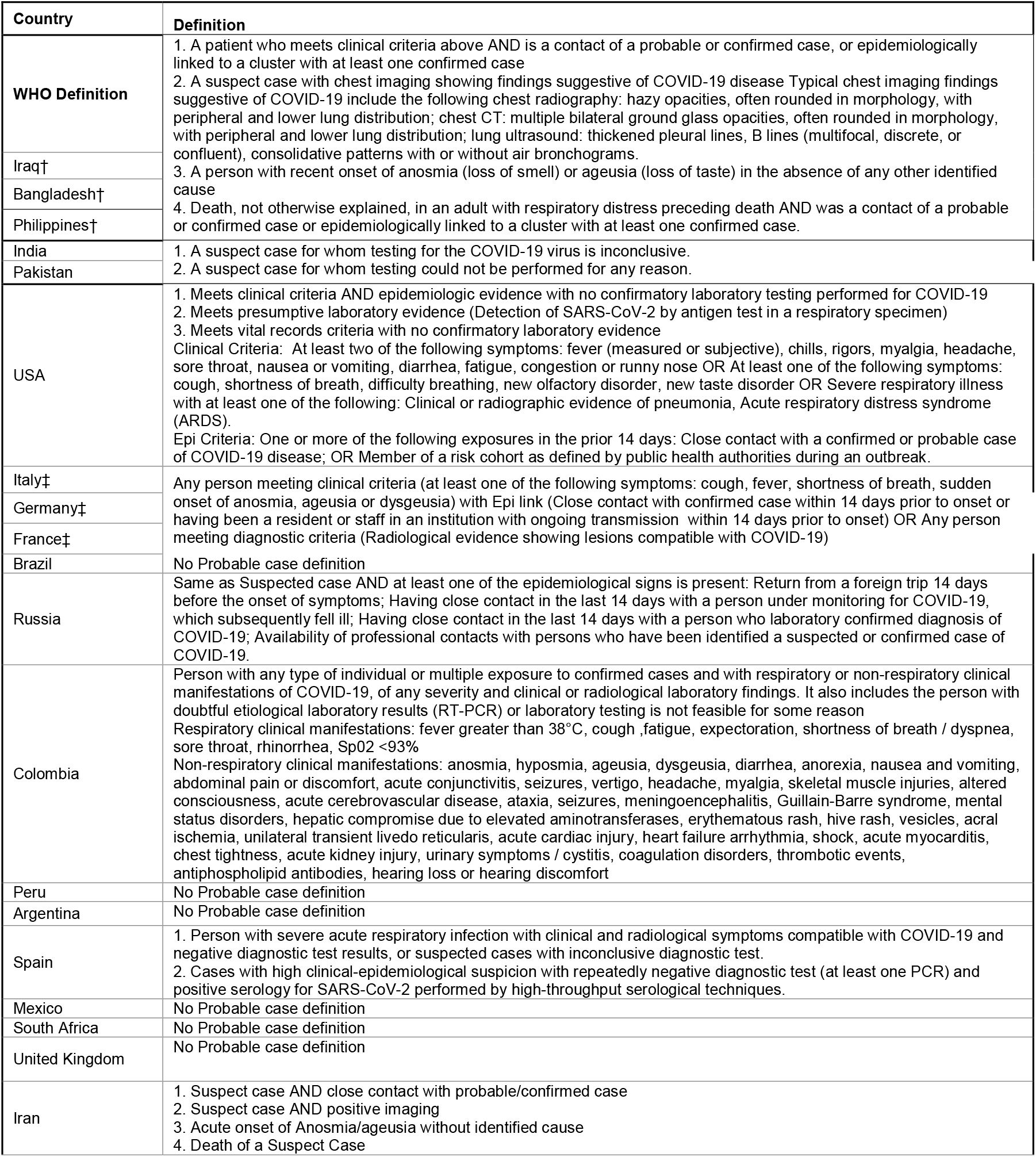
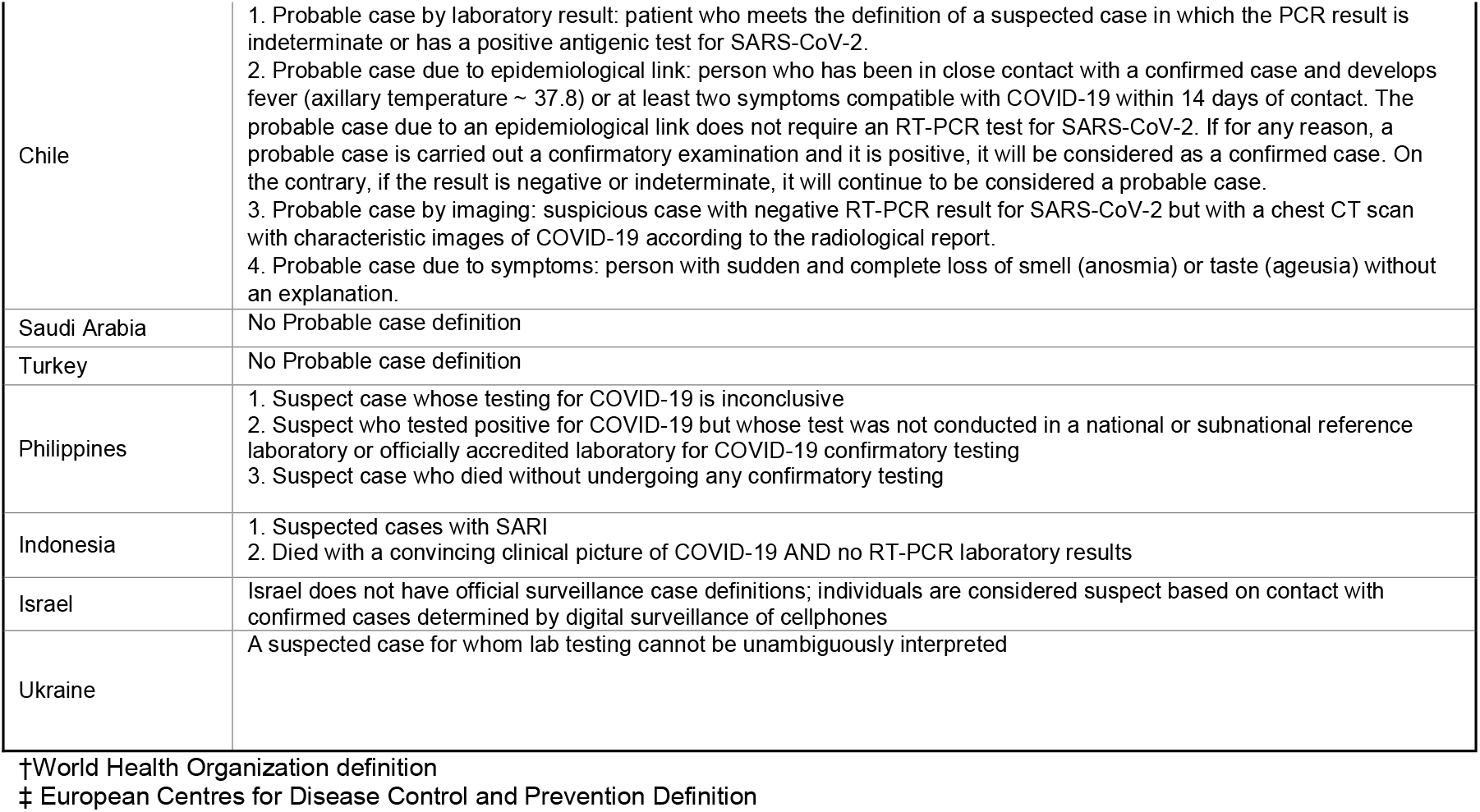

## Appendix 3.

Confirmed Definitions

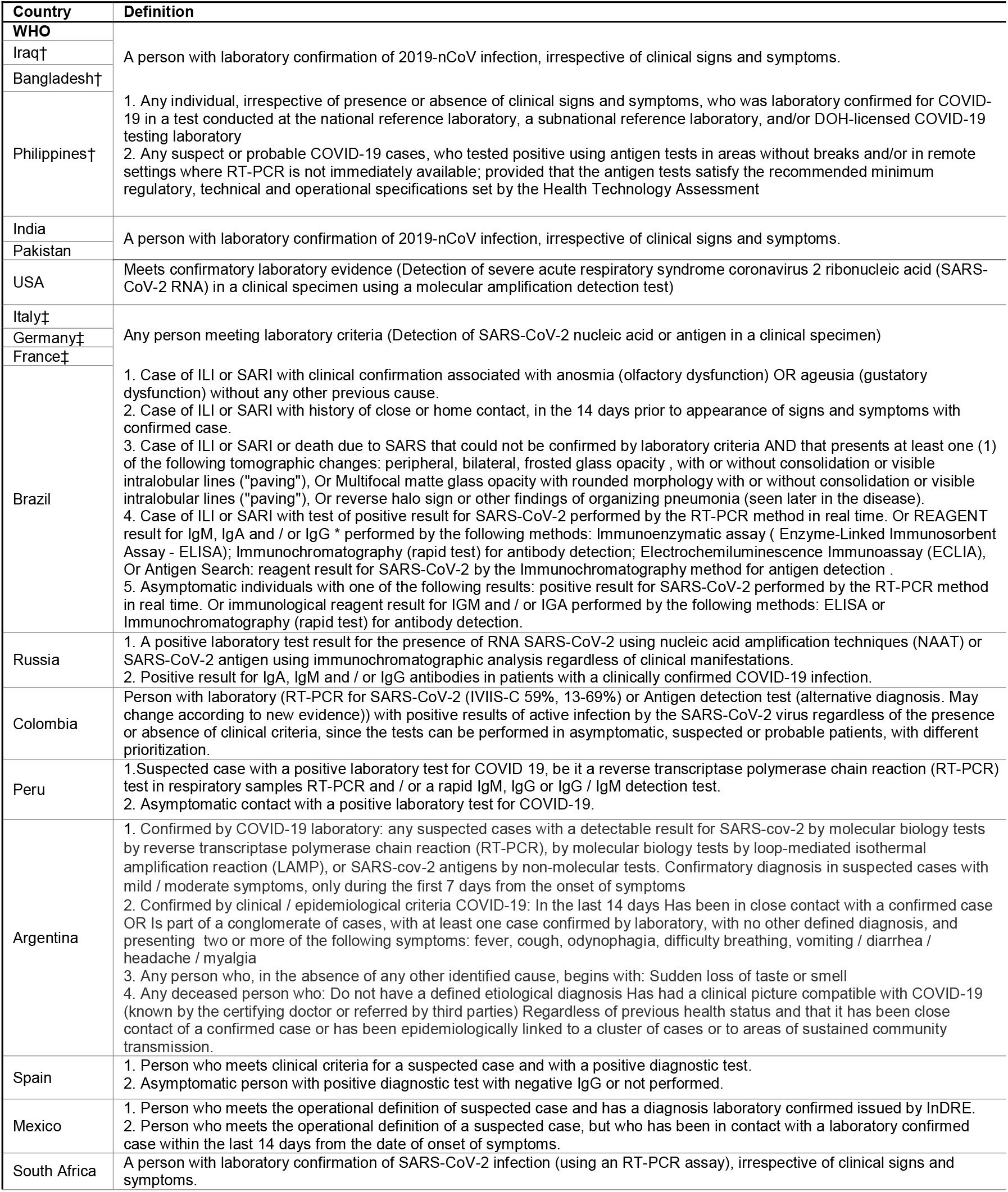
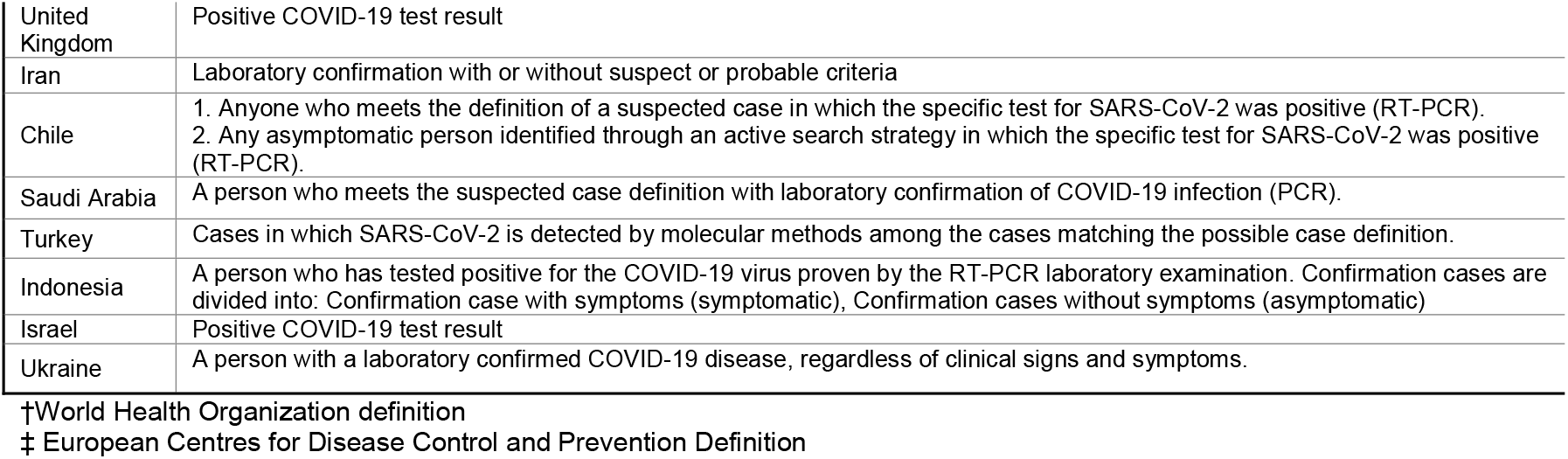

## Appendix 4.

Country Testing Policies for Asymptomatic Individuals

**Appendix 4. Country Testing Policies for Asymptomatic Individuals**
| Countries | Asymptomatic testing | EPI Criteria:<br>Confirmed<br>Contact | EPI Criteria:<br>Healthcare<br>employee |
| --- | --- | --- | --- |
| **WHO** | **Yes with EPI Criteria** | **Yes** |  |
| Iraq | Yes with EPI Criteria | Yes |  |
| Bangladesh | No |  |  |
| Philippines | Yes |  |  |
| India | Yes with EPI Criteria | Yes | Yes |
| Pakistan | Yes |  |  |
| USA | Yes |  |  |
| Italy | Yes |  |  |
| Germany | Yes with EPI Criteria | Yes | Yes |
| France | Yes |  |  |
| Brazil | Yes with EPI Criteria | Yes | Yes |
| Russia | Yes |  |  |
| Colombia | No |  |  |
| Peru | No |  |  |
| Argentina | No |  |  |
| Spain | Yes with EPI Criteria | Yes | Yes |
| Mexico | No |  |  |
| South Africa | Yes with EPI Criteria | Yes |  |
| UK | No |  | Yes |
| Iran | Yes with EPI Criteria | Yes |  |
| Chile | Yes |  |  |
| Saudi Arabia | No |  | Yes |
| Turkey | Yes with EPI Criteria | Yes | Yes |
| Indonesia | No |  |  |
| Israel | Yes |  |  |
| Ukraine | No |  |  |

